# Clinical and immunometabolic profiles of overweight and obese patients with dengue: A matched cohort study

**DOI:** 10.64898/2025.12.15.25342260

**Authors:** Nguyen Lam Vuong, Nguyet Nguyen Minh, Dong Thi Hoai Tam, Duyen Huynh Thi Le, Huy Huynh Le Anh, THuy Huynh Le Phuong, Kieu Nguyen Tan Thanh, Nguyen Thanh Van, Nguyen Thi Xuan Chau, Vi Tran Thuy, Lam Phung Khanh, Cao Thi Tam, Nguyen Thanh Phong, Nguyen Thi Hong Lan, Luong Thi Hue Tai, Nguyen Thi Cam Huong, Phan Vinh Tho, Phan Tu Qui, Huynh Trung Trieu, Nguyen Van Hao, Tran Vinh Diet, Eben Jones, Laura Rivino, Ronald B. Geskus, Ho Quang Chanh, Sophie Yacoub

## Abstract

**Background:** Obesity is a risk factor for severe dengue, but underlying mechanisms remain unclear. This study compared clinical features, outcomes, and biomarkers between overweight/obese and healthy-weight patients with dengue infection.

**Methods:** We conducted an observational matched cohort study (2019-2022) including 75 overweight/obese patients and 75 matched healthy-weight patients with dengue (by age, sex, illness phase, and inpatient ward). The primary clinical outcome was overall severity per WHO 2009 classification. Secondary outcomes included warning signs, ICU admission, fever clearance, and hospital stay. Laboratory outcomes comprised trajectories of hematology, biochemistry, and endothelial, inflammatory, and lipid biomarkers.

**Results:** The majority of patients were enrolled on day 2-3 of illness, with DENV-2 (63.3%) as the predominant serotype and secondary infections in 93.4%. Clinical signs and symptoms at enrolment and severe dengue development (6.3% vs. 9.2%) were comparable between groups. However, the overweight/obese group developed fluid accumulation more frequently (64.1% vs. 41.5%). Laboratory profiles showed consistently higher leukocyte (∼0.55-1 K/μL), neutrophil (∼0.19-0.84 K/μL), and lymphocyte counts (∼0.04-0.28 K/μL), total cholesterol (∼0.39-0.46 mmol/L), low-density lipoprotein cholesterol (∼0.36-0.48 mmol/L), triglycerides (∼0.21-0.44 log2-mmol/L), C-reactive protein (∼0.21-0.43 log10-mg/L), ferritin (∼0.12-0.15 log10-ng/mL), and leptin (∼0.89-1 log10-ng/mL) in the overweight/obese group, alongside lower high-density lipoprotein cholesterol (∼0.06-0.23 mmol/L) and adiponectin (∼0.05-0.24 log10-ng/mL). Liver transaminases were initially higher (days 1-5) in the overweight/obese group.

**Conclusions:** Overweight/obese patients with dengue infection display distinct clinical and immunometabolic profiles, particularly greater fluid accumulation, higher inflammatory markers and liver injury. These findings suggest obesity modifies the host response to dengue, and this patient group should be prioritized for upcoming immunomodulatory and other host-directed therapeutic trials.

**Author summary:** Dengue is a major global health problem, and people with obesity appear to have a higher risk of developing severe disease. However, the reasons for this increased vulnerability are still poorly understood. In this study, we compared 75 overweight or obese patients with dengue to 75 patients of healthy weight who were carefully matched by age, sex, disease stage, and hospital ward.

Although both groups had similar symptoms at enrollment and similar rates of severe dengue, the overweight/obese group developed fluid accumulation more often-a key sign of clinical deterioration. They also showed distinct laboratory patterns throughout their illness. Overweight/obese patients consistently had higher levels of white blood cells, inflammatory markers, liver enzymes, and lipid measures, as well as higher leptin and lower adiponectin, two hormones linked to metabolic health.

These findings suggest that excess body weight alters the body’s immune and metabolic response to dengue infection, even when outward symptoms appear comparable. Understanding these differences is important because it can help clinicians identify patients at higher risk and guide the development of new treatments. Our results support prioritizing overweight and obese individuals for future trials of immunomodulatory or other host-directed therapies for dengue.

## Introduction

Dengue continues to be a major public health burden globally, with a dramatic growth in disease incidence in recent decades [1]. While most symptomatic dengue cases resolve after 1 week, approximately 5-10% of hospitalized patients develop severe manifestations during the critical phase (day 4-6 of illness) [2]. The progression to severe dengue has been found to be associated with multiple factors, including certain comorbidities, such as obesity [3–6].

Many low-to-middle-income countries in the dengue endemic areas have seen a large increase in the number of overweight/obese children and young adults in the last two decades. The underlying mechanisms of the association between obesity and dengue severity remain speculative, but may be linked to the chronic low-grade inflammation that occurs in obesity, with adipocytes producing pro-inflammatory cytokines, as well as adipokines (e.g., adiponectin, resistin, leptin) which have immunomodulatory effects [7]. Leptin is the most important pathophysiological mediator of inflammation [7–9]. Obesity-induced leptin and resistin are associated with platelet dysfunction, vascular inflammation, myocardial injury, and endothelial dysfunction [10, 11], as well as capillary hyper-permeability [12, 13], which could contribute to dengue pathogenesis. Several other immunological abnormalities have been associated with obesity, such as the impaired antibody and T cell responses and defective Natural Killer (NK) cell activity which can impair host defenses against viral diseases [14–16]. The underlying mechanisms for the increased risk of severe complications in obese patients with dengue needs further research.

We hypothesized that obesity modulates dengue disease outcomes through increased lipid-inflammatory mediators, adipokines and immune dysfunction. We aimed to investigate some of the underlying mechanisms by comparing clinical manifestations, clinical outcomes, and specific biomarkers levels between healthy-weight and overweight/obese dengue patients. In this paper, we report the data related to comparative clinical evolution of dengue and kinetics of endothelial, inflammatory and lipid markers between participants with obesity and their matched healthy-weight controls, who were all hospitalized with lab-confirmed dengue.

## Methods

### Study design and participants

This was an observational matched cohort study for overweight/obese adolescent and young adult patients with dengue and a control group consisting of healthy-weight dengue patients admitted to the Hospital for Tropical Diseases (HTD) in Ho Chi Minh city (HCMC), Vietnam, between May 2019 and June 2022. The study was approved by the Ethics Committee of the HTD (CS/BND/19/07) and the Oxford Tropical Research Ethics Committee (OxTREC 5-19). Written informed consent was obtained from all adult participants (≥18 years old) or the parents (children under 12 years old). Those between 12 and 18 years of age were asked to sign an assent form and the parents provided written consent to give permission for their child to participate in the study.

Overweight/obese patients were defined by the body mass index (BMI) of >25 kg/m^2^ in patients >19 years of age or the BMI-for-age of >1 standard deviation (SD) in those ≤19 years of age. Healthy-weight patients were defined by the BMI between 17 and 22 kg/m^2^ for adults or in normal range by age for children (between -1 and 0 SD) [17].

Patients were identified and approached when they were admitted to the dengue wards of the HTD, including those referred from any other hospitals for admission or assessment of suspected dengue. The inclusion criteria were: (i) a confirmed dengue diagnosis with a positive NS1 rapid test, (ii) males or females from 10 to 30 years of age, (iii) ≤4 days (≤96 hours) since fever onset amongst general ward patients or ≤5 days (≤120 hours) since fever onset amongst those admitted to intensive care units (ICUs), (iv) either overweight/obese or healthy-weight based on BMI as described previously, and (v) agreed to attend a final follow-up (FU) visit at around day 21-28 of illness. We excluded pregnant women; patients with signs or symptoms suggesting other acute infectious diseases; or those who were unlikely to attend the FU visit based on the judgement of study physicians.

Each case in the healthy-weight group was matched with each enrolled overweight/obese patient by four factors: (i) age group (10-15; 16-20; 21-25; and 26-30), (ii) sex (male and female), (iii) illness phase (febrile phase [fever days 1-3] and critical phase [fever days 4-5]), and (iv) inpatient ward at enrolment (general ward and ICU).

### Dengue diagnostics

Dengue viral serotyping and viremia levels were performed at enrolment using the reverse transcription-polymerase chain reaction (RT-PCR) assay that has been described elsewhere [18]. Serology testing was done on paired samples taken at the acute and convalescence phases of the illness, using the IgM and IgG antibody capture ELISAs.

Dengue was laboratory-confirmed when patients had either: (i) a positive PCR assay; or (ii) a positive NS1 rapid test; or (iii) a seroconversion of MAC-ELISA on paired samples. Classification of primary and secondary dengue was based on the presence of IgG on paired samples. A probable primary infection was defined by two negative/equivocal IgG results on separate samples taken at least two days apart within the first ten days of symptom onset, with at least one sample during the convalescent phase (days 6-10). A probable secondary infection was defined by at least one positive IgG result during the first ten days. Cases without time-appropriate IgG results were classified as indeterminate.

### Routine clinical and laboratory assessments and blood sampling

The progression of clinical signs and symptoms in all patients were assessed daily for up to five days or until discharge (whichever came first) with a standardized case report form. General management during the hospitalization was at the discretion of the attending physicians.

During hospitalization, full blood count (FBC), glucose and lactate point-of-care (POC) tests and research blood sampling (for dengue diagnostics, biomarker investigations and immunology studies) were performed daily. Biochemistry consisting of liver transaminases (aspartate aminotransferase [AST] and alanine aminotransferase [ALT]), creatinine, albumin, C-reactive protein (CRP) and lipid profile (total cholesterol, high density lipoprotein cholesterol [HDL-c], low density lipoprotein cholesterol [LDL-c], and triglycerides) were tested on every other day.

All patients were asked to return for a final follow-up visit at around 3-4 weeks after fever onset, when their health was assessed together with the final blood samples for FBC, biochemistry tests and research (serology diagnostic, immunology studies).

### Clinical outcomes

The primary clinical outcome was disease severity, which was categorized according to the WHO 2009 classification [2]– dengue, dengue with warning signs, and severe dengue – based on all available data. Severe dengue was defined by severe plasma leakage (dengue shock syndrome [DSS] or respiratory distress due to plasma leak), severe bleeding, or severe major organ involvement (including central nervous system, liver, kidney, and cardiac involvement).

Secondary clinical outcomes were also analyzed and included specific warning signs, ICU admission, fever clearance time, and length of hospital stay. Laboratory outcomes were the trajectory of the full blood count, biochemistry, and biomarkers, as well as platelet nadir and maximum haematocrit change from baseline (i.e., minimum haematocrit within illness day 1-3).

### Biomarker outcomes

Concentrations of endothelial, inflammatory, and lipid biomarkers were measured on study days 1, 2, 3, 5, and at the follow-up visit. Adiponectin, leptin, and ferritin were quantified using ELISA kits (Arigo Biolaboratories, Zhubei, Taiwan; and Bio-Techne Corporation, Minnesota, USA). The remaining biomarkers were analyzed using a multiplex premixed magnetic bead-based assay on a Luminex Flexmap3D platform (R&D Systems, Bio-Techne Corporation, Minnesota, USA). These included interleukin (IL)-6, IL-8, IL-10, IL-18, IL-1 receptor antagonist (IL-1RA), angiopoietin-2, syndecan-1, proprotein convertase subtilisin/kexin type 9 (PCSK9), resistin, C-X-C motif chemokine ligand 10 (CXCL-10), vascular cell adhesion molecule 1 (VCAM-1), myeloperoxidase (MPO), and growth/differentiation factor 15 (GDF-15). All assays were performed according to the manufacturers’ instructions.

The NS1 levels were quantified using an in-house quantitative sandwich enzyme immunoassay. Dengue NS1 in patients’ plasma was captured by a highly specific dengue mouse monoclonal antibody (7e11) and then this antigen-antibody complex was detected using a biotinylated highly specific mouse antibody monoclonal (2B7).

### Statistical analysis

A sample size of 75 patients per group (150 patients in total) was determined based on clinical judgement and feasibility considerations.

All baseline characteristics and clinical endpoints were summarized by the two groups using median and 25^th^; 75^th^ percentiles for continuous variables, and the number of patients and percentage for categorical variables. Values of continuous variables with skewed distribution such as biomarker levels were transformed to get a nearly normal distribution. Because of the matching design, the Wilcoxon signed-rank test and McNemar’s test were used to assess differences in clinical symptoms and outcomes between groups.

Linear mixed-effects models were utilized for the trajectory of the routine laboratory test results, and endothelial, inflammatory, and lipid biomarker levels. The fixed effects included the main exposure (overweight/obesity vs. healthy-weight), illness day of measurement, and their interaction. Random effects included intercepts for patients and matching. Illness day was treated as a continuous variable during the first 10 days in models for routine laboratory tests, but as a categorical variable (days 1-2, 3-4, 5-6, 7-8, and follow-up days 19-40) in models for biomarkers, owing to the limited number of biomarker measurements (up to three timepoints during the acute phase and once at follow-up). Restricted cubic splines with three knots at 2, 4, and 7 were applied for illness day in the models for routine laboratory tests. The routine laboratory test results at the follow-up visit were analyzed separately using linear regression models with covariates main exposure, illness day of measurement (splines with knots at 20, 25, and 30), and their interaction. The models for IL-6, IL-8, IL-10, and NS1 levels were allowed for left censored values because these biomarkers had values below the limit of detection. Results from the models were mainly reported by visualization of the predicted values by the two groups.

All the analyses were performed using the statistical software R version 4.1.3.

## Results

A total of 135 overweight/obese patients and 131 healthy-weight patients were screened, and 75 were enrolled in each group. Each group included 65 patients from general dengue wards and 10 from ICUs. One patient in the overweight/obese group (at a general ward) withdrew from the study during the follow-up period (**Figure 1**).

**Figure 1.**
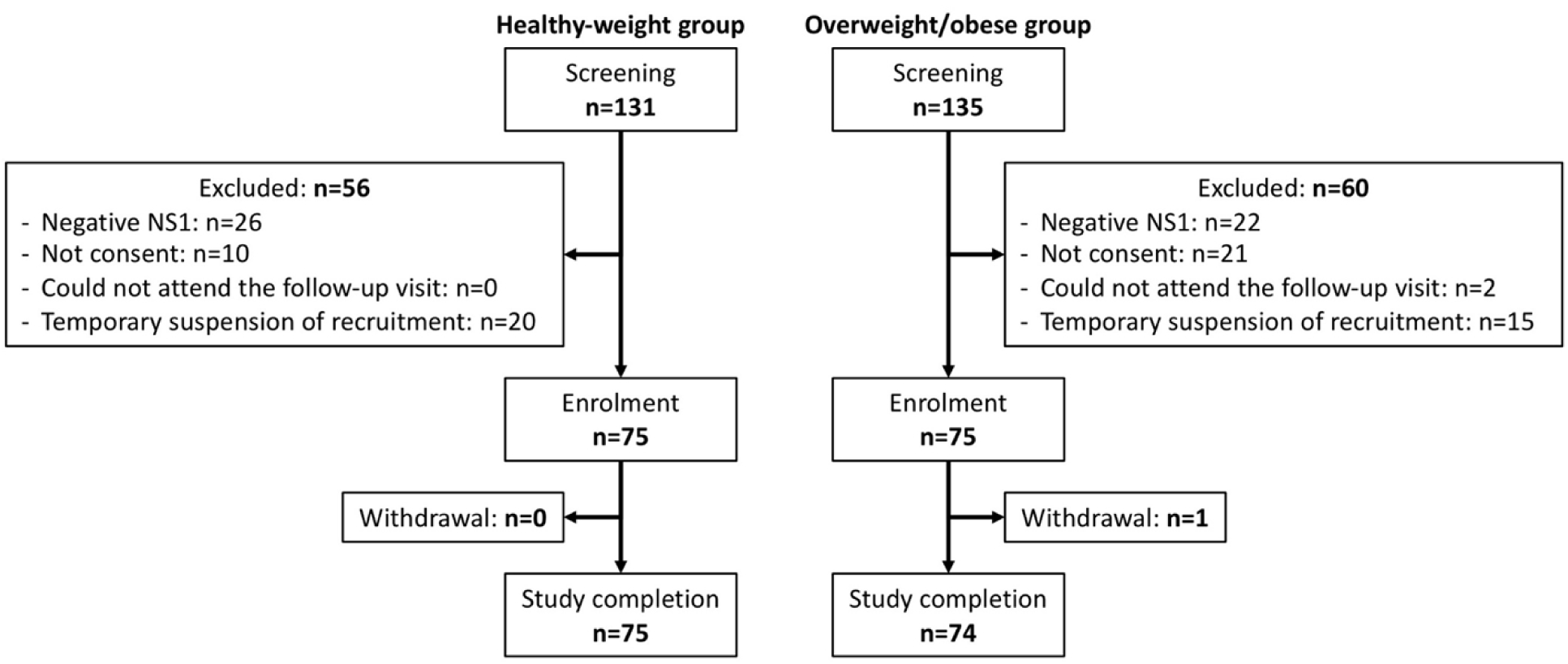
Study flowchart

### Baseline characteristics and clinical signs and symptoms at enrolment

Baseline characteristics including age, sex, and inpatient ward and illness day at enrolment were balanced between the two groups since they were the matching variables. The median age was 15.5 years for patients enrolled from general wards and 13 years for those from ICUs. Males were predominant, accounting for 78.7%. At enrolment, most patients in general wards were in the febrile phase (illness day 2 or 3), whereas all ICU patients were at illness day 4 or 5 (**Table 1**). Serotype and immune status were balanced between groups, with DENV-2 being the predominant serotype (61.3% in the healthy-weight group and 65.3% in the overweight/obese group). Most patients were classified as secondary infection (90.7% and 96% respectively). Viremia and NS1 levels were comparable between the two groups at enrolment.

**Table 1.**
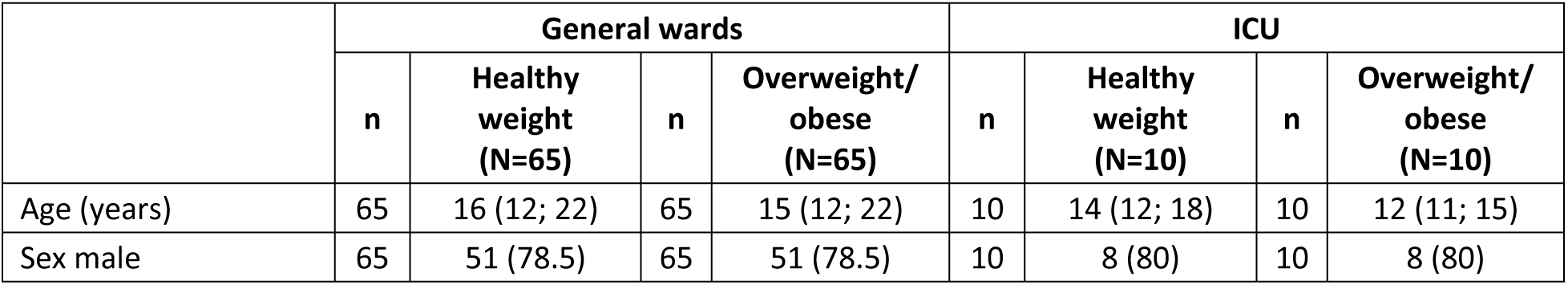
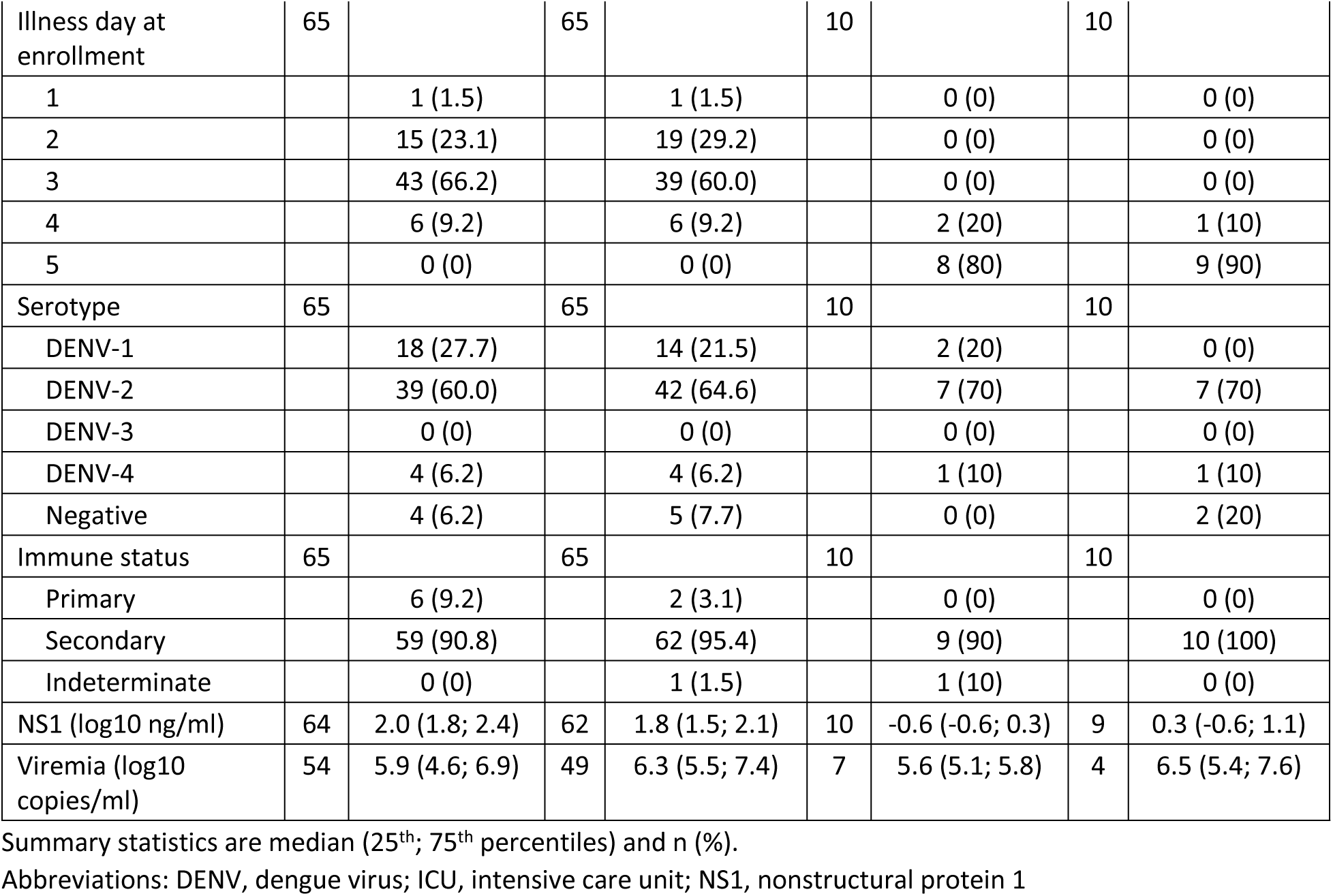
Baseline characteristics

We observed that the overweight/obese group had higher percentages of nausea, feeling dizzy/faint, vomiting, coughing, abdominal pain, liver enlargement, and clinical pleural effusion at enrolment (**Table 2**). However, percentage of obesity patients hospitalised with at least one warning signs was lower than that of healthy-weight patients (54.7% vs. 64%, respectively). The two groups had relatively similar percentages of other clinical signs and symptoms at enrolment.

**Table 2.** Clinical signs and symptoms at enrolment

|  | General wards |  |  | ICU |  |  |
| --- | --- | --- | --- | --- | --- | --- |
|  | Healthy weight (N=65) | Overweight/obese (N=65) | p-value | Healthy weight (N=10) | Overweight/obese (N=10) | p-value |
| Any warning sign | 38 (58.5) | 31 (47.7) | 0.248 | 10 (100) | 10 (100) | - |
| Headache and/or muscle/joint pain | 63 (96.9) | 63 (96.9) | 1 | 10 (100) | 10 (100) | - |
| Bleeding | 40 (61.5) | 38 (58.5) | 0.838 | 10 (100) | 10 (100) | - |
|  | Healthy weight (N=65) | Overweight/obese (N=65) | p-value | Healthy weight (N=10) | Overweight/obese (N=10) | p-value |
| - Bruising/Petechia | 38 (58.5) | 34 (52.3) | 0.556 | 10 (100) | 10 (100) | - |
| - Gum/nose bleeding | 4 (6.2) | 6 (9.2) | 0.724 | 1 (10) | 2 (20) | 1 |
| - Vaginal bleeding | 2/14 (14.3) | 1/14 (7.1) | 1 | 0/2 (0) | 2/2 (100) | - |
| - Hematemesis/melaena/hematuria | 2 (3.1) | 1 (1.5) | 1 | 0 (0) | 0 (0) | - |
| - Other bleeding | 0 (0) | 0 (0) | - | 0 (0) | 0 (0) | - |
| Nausea | 42 (64.6) | 47 (72.3) | 0.499 | 7 (70) | 8 (80) | 1 |
| Feeling dizzy/faint | 39 (60.0) | 44 (67.7) | 0.458 | 8 (80) | 9 (90) | 1 |
| Vomiting | 31 (47.7) | 32 (49.2) | 1 | 7 (70) | 8 (80) | 1 |
| Coughing | 28 (43.1) | 40 (61.5) | 0.059 | 6 (60) | 4 (40) | 0.617 |
| Abdominal pain | 22 (33.8) | 26 (40.0) | 0.556 | 6 (60) | 8 (80) | 0.683 |
| Abdominal tenderness | 17 (26.2) | 9 (13.8) | 0.136 | 8 (80) | 9 (90) | 1 |
| Diarrhoea | 23 (35.4) | 24 (36.9) | 1 | 2 (20) | 1 (10) | 1 |
| Skin rash | 18 (27.7) | 18 (27.7) | 1 | 3 (30) | 4 (40) | 1 |
| Liver enlargement | 0 (0) | 3 (4.6) | - | 9 (90) | 9 (90) | 1 |
| Clinical pleural effusion | 0 (0) | 0 (0) | - | 6 (60) | 9 (90) | 0.248 |
| Ascites | 0 (0) | 0 (0) | - | 3 (30) | 2 (20) | 1 |
| Lethargy | 2 (3.1) | 3 (4.6) | 1 | 0 (0) | 2 (20) | - |
| Jaundice | 0 (0) | 0 (0) | - | 0 (0) | 0 (0) | - |
| Spleen enlargement | 0 (0) | 0 (0) | - | 0 (0) | 0 (0) | - |
| Restlessness | 0 (0) | 0 (0) | - | 0 (0) | 0 (0) | - |
| Neurological abnormality | 0 (0) | 0 (0) | - | 0 (0) | 0 (0) | - |

Summary statistics are n (%).
Abbreviations: ICU, intensive care unit

### Clinical outcomes

The rates of severe clinical outcomes were similar in the two groups. Severe dengue occurred in 6.3% and 9.2% of the overweight/obese and healthy-weight groups enrolled from general wards, mostly due to DSS (6.3% and 7.7%, respectively). Severe organ impairment occurred in two patients in the healthy-weight group (one enrolled from general ward and one from ICU), both had severe liver involvement. None had severe bleeding (**Table 3**).

**Table 3.** Clinical outcomes

|  | General wards |  |  |  |  | ICU |  |  |  |  |
| --- | --- | --- | --- | --- | --- | --- | --- | --- | --- | --- |
|  | n | Healthy weight (N=65) | n | Overweight / obese (N=65) | p-value | n | Healthy weight (N=10) | n | Overweight / obese (N=10) | p-value |
| **Dengue clinical outcomes** |  |  |  |  |  |  |  |  |  |  |
| WHO 2009 classification | 65 |  | 64 |  | 0.853 | 10 |  | 10 |  | - |
| Dengue |  | 9 (13.8) |  | 7 (10.9) |  |  | 0 (0) |  | 0 (0) |  |
| Dengue with warning signs |  | 50 (76.9) |  | 53 (82.8) |  |  | 0 (0) |  | 0 (0) |  |
| Severe dengue |  | 6 (9.2) |  | 4 (6.3) |  |  | 10 (100) |  | 10 (100) |  |
| Dengue shock syndrome | 65 | 5 (7.7) | 64 | 4 (6.3) | 1 | 10 | 10 (100) | 10 | 10 (100) | - |
| Recurrent shock | 5 | 0 (0) | 4 | 1 (25.0) | - | 10 | 3 (30) | 10 | 2 (20) | 1 |
| Prolonged shock | 5 | 1 (20.0) | 4 | 0 (0) | - | 10 | 0 (0) | 10 | 1 (10) | - |
| Respiratory distress | 65 | 0 (0) | 64 | 0 (0) | - | 10 | 0 (0) | 10 | 1 (10) | - |
| Severe bleeding | 65 | 0 (0) | 64 | 0 (0) | - | 10 | 0 (0) | 10 | 0 (0) | - |
| Severe liver involvement | 65 | 1 (1.5) | 64 | 0 (0) | - | 10 | 1 (10) | 10 | 0 (0) | - |
| CNS involvement | 65 | 0 (0) | 64 | 0 (0) | - | 10 | 0 (0) | 10 | 0 (0) | - |
| Other major organ involvement | 65 | 0 (0) | 64 | 0 (0) | - | 10 | 0 (0) | 10 | 0 (0) | - |
| **Specific warning signs according to WHO 2009 classification** |  |  |  |  |  |  |  |  |  |  |
| Any warning sign | 65 | 57 (87.7) | 64 | 57 (89.1) | 1 | 10 | 10 (100) | 10 | 10 (100) | - |
| Abdominal pain | 65 | 37 (56.9) | 64 | 29 (45.3) | 0.281 | 10 | 7 (70) | 10 | 10 (100) | - |
| Fluid accumulation | 65 | 27 (41.5) | 64 | 41 (64.1) | 0.012 | 10 | 10 (100) | 10 | 10 (100) | - |
| Increase in HCT concurrent with rapid decrease in PLT | 65 | 26 (40.0) | 64 | 24 (37.5) | 1 | 10 | 10 (100) | 10 | 10 (100) | - |
| Mucosal bleeding | 65 | 31 (47.7) | 64 | 25 (39.1) | 0.486 | 10 | 3 (30) | 10 | 4 (40) | 1 |
| Persistent vomiting | 65 | 17 (26.2) | 64 | 9 (14.1) | 0.118 | 10 | 1 (10) | 10 | 1 (10) | 1 |
| Liver enlargement > 2cm | 65 | 11 (16.9) | 64 | 12 (18.8) | 1 | 10 | 7 (70) | 10 | 7 (70) | 1 |
| AST and/or ALT > 400 U/L | 65 | 4 (6.2) | 64 | 6 (9.4) | 0.752 | 10 | 1 (10) | 10 | 0 (0) | - |
| Lethargy | 65 | 2 (3.1) | 64 | 4 (6.3) | 0.683 | 10 | 0 (0) | 10 | 2 (20) | - |
| Restlessness | 65 | 1 (1.5) | 64 | 0 (0) | - | 10 | 1 (10) | 10 | 0 (0) | - |
| ICU admission | 65 | 4 (6.2) | 65 | 2 (3.1) | 0.683 | 10 | 10 (100) | 10 | 10 (100) | - |
| ICU length of stay (hours) | 4 | 56 (41; 68) | 2 | 65 (53; 78) | - | 10 | 43 (26; 62) | 10 | 49 (33; 71) | 0.570 |
| Fever clearance time (hours) | 65 | 111 (92; 132) | 64 | 112 (102; 127) | 0.738 | 10 | 103 (94; 108) | 10 | 102 (94; 105) | 0.492 |
| Hospital length of stay (days) | 65 | 7 (7; 8) | 63 | 7 (7; 8) | 0.467 | 10 | 6 (5; 6) | 10 | 7 (6; 7) | 0.205 |
| Platelet nadir (K/uL) | 65 | 44 (18; 72) | 65 | 44 (19; 81) | 0.810 | 10 | 11 (9; 14) | 10 | 15 (8; 21) | 0.575 |
| Max haematocrit change (%) | 65 | 14 (7; 19) | 64 | 12 (7; 17) | 0.241 | 4 | 5 (-2; 19) | 8 | 5 (-3; 14) | 0.625 |

Summary statistics are median (25<sup>th</sup>; 75<sup>th</sup> percentiles) and n (%).
Abbreviations: ALT, alanine aminotransferase; AST, aspartate aminotransferase; CNS, central nervous system; HCT, haematocrit; ICU, intensive care unit; PLT, platelet; WHO, World Health Organization

Most patients developed more warning signs during their hospitalization (89.1% and 87.7% in the overweight/obese and healthy-weight group enrolled from general wards, respectively). Among those, abdominal pain was the most frequent, followed by fluid accumulation, increase in haematocrit concurrent with rapid decrease in platelet count, and mucosal bleeding. Among patients enrolled from general wards, the overweight/obese group had significantly higher proportion of fluid accumulation compared to the healthy-weight group (64.1% vs. 41.5%) (**Table 3**). There were no significant differences between the two groups regarding other warning signs. Other outcomes were also similar in the two groups, including ICU admission, length of ICU stay, fever clearance time, length of hospital stay, platelet nadir, and max haemoconcentration (**Table 3**).

### Laboratory tests and biomarkers

There were several differences in the routine laboratory tests between the two patient groups (**Figure 2**, **Table S1**). Regarding full blood count, the overweight/obese group had persistently higher white cell counts (difference in estimated mean: 0.55 to 1 K/μL), neutrophil counts (difference in estimated mean: 0.19 to 0.84 K/μL), and lymphocyte counts (difference in estimated mean: 0.04 to 0.28 K/μL) during the course of illness and at recovery, whereas platelet counts and haematocrit were similar in the two groups.

**Figure 2.**
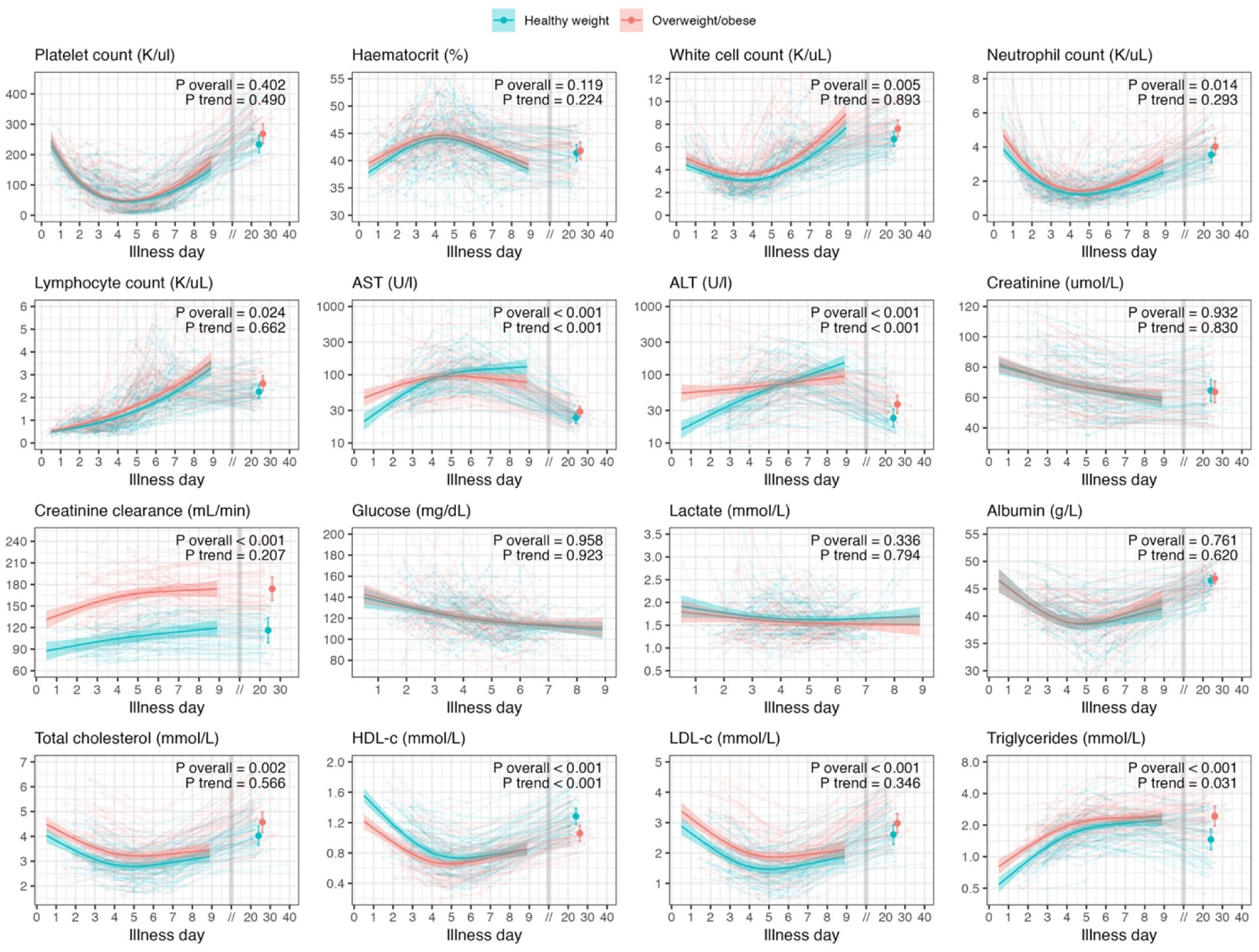
Full blood count and biochemistry

Thick-coloured lines and coloured regions represent predicted values and 95% CIs from the linear mixed-effects models. Thick-coloured dots and whiskers represent predicted values and 95% CIs from the linear regression models at the follow-up period after day 18. Light-coloured lines represent the individual trajectory of the parameters. P values are derived from the linear mixed-effects model. P overall represents the overall effect of overweight/obesity, and P trend compares the trend over time between the two groups.

Abbreviations: ALT, alanine aminotransferase; AST, aspartate aminotransferase; CI, confidence interval; HDL-c, high density lipoprotein cholesterol; LDL-c, low density lipoprotein cholesterol Transaminase levels appeared to increase earlier in the overweight/obese group compared with the healthy-weight group, with higher levels observed during the first 4-5 days of illness (difference in estimated mean AST and ALT: 10 to 25 U/L and 18 to 38 U/L, respectively). From day 5 to 9, however, the overweight/obese group had lower transaminase levels (difference in estimated mean AST and ALT: 10 to 54 U/L and 4 to 55 U/L, respectively) (**Figure 2**). During hospitalization, two patients developed severe liver involvement (AST >1000 U/L), both of whom were in the healthy-weight group.

The two groups showed similar levels and kinetics of creatinine, glucose, lactate, and albumin. Regarding lipid profile, the overweight/obese group had higher total cholesterol, LDL-c, and triglycerides, and lower HDL-c during the illness course. In both groups, concentrations of all cholesterol components decreased during the acute dengue episode, reaching a nadir around day 5 of illness, while triglyceride levels increased from fever onset until discharge (**Figure 2**).

With respect to endothelial, inflammatory, and lipid biomarkers, the overweight/obese group consistently showed higher levels of CRP (difference in estimated mean: 1 to 6.6 mg/L), ferritin (difference in estimated mean: 52 to 114 ng/mL), and leptin (difference in estimated mean: 7 to 13 ng/mL), but lower levels of adiponectin (difference in estimated mean: 615 to 1295 ng/mL) (**Figure 3**, **Table S2**). For IL-6, the two groups showed similar levels on illness days 3-6, but the overweight/obese group had significantly higher levels on days 7-8 (difference in estimated mean: 15 ng/mL). For other biomarkers—including IL-8, IL-10, angiopoietin-2, syndecan-1, IL-1RA, resistin, CXCL-10, VCAM-1, MPO, GDF-15—both groups demonstrated similar levels and trends.

**Figure 3.**
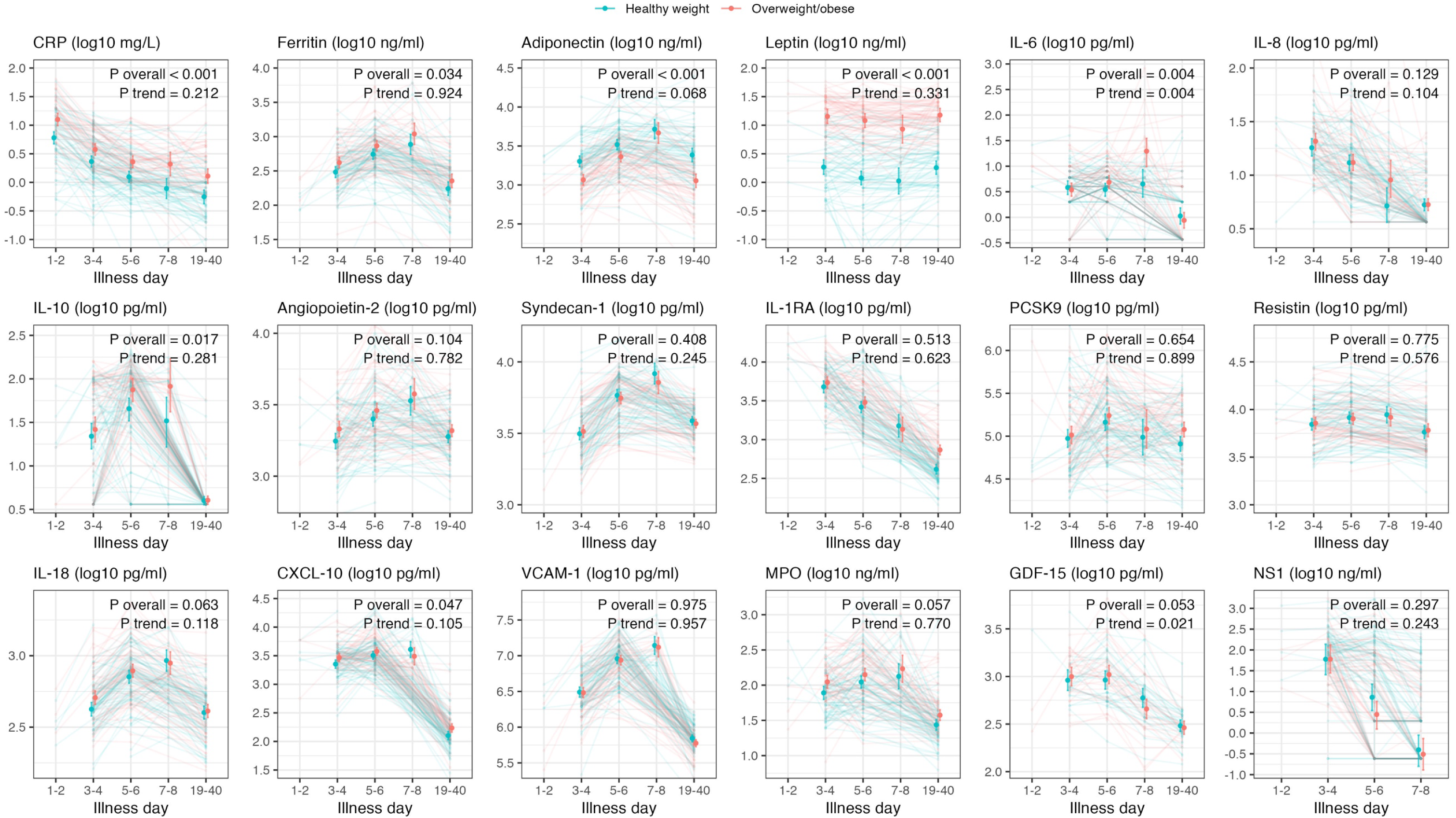
Endothelial, inflammatory, and lipid biomarkers

Thick-coloured dots and whiskers represent predicted values and 95% CIs from the linear mixed-effects models. Light-coloured lines represent the individual trajectory of the parameters. P values are derived from the linear mixed-effects model. P overall represents the overall effect of overweight/obesity, and P trend compares the trend over time between the two groups. Abbreviations: CI, confidence interval; CRP, C-reactive protein; CXCL-10 (IP-10), CXC motif chemokine ligand 10; FU, follow-up; GDF-15, growth/differentiation factor 15; IL-10, interleukin-10; IL-18, interleukin-18; IL-1RA, interleukin-1 receptor antagonist; IL-8, interleukin-8; MPO, myeloperoxidase; NS1, nonstructural protein 1; PCSK9, proprotein convertase subtilisin/kexin type 9; VCAM-1, vascular cell adhesion molecule 1

## Discussion

This matched cohort study is one of the first to comprehensively evaluate the clinical manifestations and biomarker trajectories in hospitalized dengue patients stratified by nutritional status. We observed that patients with overweight/obesity presented with significantly higher frequencies of fluid accumulation—a symptom included in the WHO 2009 dengue warning signs. However, the incidence of progression to severe dengue, mostly DSS, was comparable between the overweight/obese group and their healthy-weight counterparts.

The overweight/obese group consistently exhibited elevated levels of white blood cell counts (including both neutrophil and lymphocyte), inflammatory (CRP, ferritin) and metabolic (leptin) biomarkers, as well as altered lipid profiles (higher LDL-c, triglycerides, and lower HDL-c). CRP is an inflammatory marker that is associated with higher risk of progression to severe dengue and may contribute to dengue pathogenesis by promoting endothelial dysfunction [19, 20]. Elevated leptin and low adiponectin are characteristic in obesity [21]. Altered adipokine levels are thought to mediate excessive host inflammation in obesity, and can also contribute to endothelial and platelet dysfunction [20]. The immunometabolic characteristics observed in the patients with obesity may therefore plausibly mechanistically underpin the higher rate of fluid accumulation in this population. These findings support existing hypotheses that obesity, through chronic low-grade inflammation and adipokine dysregulation, may prime the host immune response, potentially affecting endothelial integrity and vascular permeability during dengue infection. This is consistent with a previous study reporting that altered T-cell and NK-cell responses, which were more pronounced in overweight/obese patients compared with healthy-weight patients, were early signatures in those who later developed severe dengue [22]. Interestingly, while liver enzyme levels (AST, ALT) were higher at baseline in the overweight/obese group, their peak levels were lower and occurred earlier, suggesting differential hepatic involvement or kinetics of liver injury.

Previous studies have reported inconsistent associations between obesity and dengue infection and severity. Some paediatric studies in Latin America and Southeast Asia suggested increased risks of dengue infection, hospitalization, DSS or fluid accumulation in obese children [6, 23–27], while other adult cohorts have not demonstrated significant associations with severe outcomes [28]. Our study contributes to this literature by integrating dynamic biomarker data and revealing that although inflammatory responses differ by weight status, these do not necessarily predict worse clinical outcomes in a hospitalized population. This finding echoes research in influenza and COVID-19, where obesity was linked to altered immune responses and delayed viral clearance but not always to increased mortality.

Despite these differences in the host response, the comparable clinical outcomes may reflect the limitations of a small sample size. Furthermore, the relatively young age and overall mild-to-moderate disease burden in our cohort may attenuate potential obesity-related risks. A longitudinal design with larger sample sizes and inclusion of younger children, older adults, and those with comorbidities may be necessary to robustly evaluate the independent contribution of obesity to severe dengue outcomes. At present, there are no therapeutic options licensed for use in dengue [29]. Ongoing clinical studies are evaluating host-directed therapies that target the pathological hyperinflammation characteristic of severe dengue [30], and the distinct inflammatory and metabolic features amongst the obesity group hints that these patients may particularly benefit from this therapeutic approach.

These results highlight the importance of considering host metabolic status when interpreting clinical and laboratory features of dengue. While overweight/obese individuals may present with distinct clinical features and show evidence of higher systemic inflammation, clinical management should still rely on established criteria for fluid management and monitoring. The pathophysiological basis for the observed differences—especially the interplay of adipokines, immune dysregulation, and endothelial biomarkers—warrants further mechanistic studies, possibly including transcriptomic or proteomic profiling.

Strengths of our study include the prospective design, rigorous matching, and comprehensive biomarker analysis. Limitations include the single-center setting, modest sample size, and limited generalizability to older or comorbid populations. Moreover, although viremia and NS1 levels were measured, their association with immune response and clinical outcomes requires further exploration. Future studies should also assess viral kinetics and immunogenetic profiles to better elucidate host-pathogen interactions.

In conclusion, overweight and obese patients with dengue exhibit distinct clinical and immunometabolic profiles compared to healthy-weight peers, characterized by more frequent fluid accumulation and elevated inflammatory biomarkers. However, these differences did not translate into increased rates of severe dengue in our cohort. Our findings suggest that obesity modulates the host response to dengue infection underscoring the need for further mechanistic studies and risk stratification tools that integrate metabolic and immunologic parameters. The hyperinflammatory phenotype observed in overweight/obese patients with dengue means these patients should be prioritized for upcoming immunomodulatory therapeutic trials.

## Statements

### Competing interests

- LR: consulting for Novartis around immunomodulatory treatment for dengue, and receiving a grant from the Academy of Medical Sciences (Springboard Award) to perform immunology work on samples collected from patients included in this study. The results from our immunology study are included in the manuscript: Gregorova et al Nat Commun 2025.
- All other authors have no competing interests to declare.

### Financial disclosure

This study was funded by Wellcome Trust (Grant number B9R05430, to SY). The funders had no role in study design, data collection and analysis, decision to publish, or preparation of the manuscript.

## Data Availability

Data will be published in Oxford repository.

## Acknowledgements

None

## Author contributions

- NLV: formal analysis, methodology, writing – original draft preparation
- NNM: conceptualization, data curation, investigation, methodology, project administration, writing – review & editing
- DTHT: conceptualization, data curation, investigation, methodology, writing – review & editing
- DHTL: investigation, writing – review & editing
- HHLA: investigation, writing – review & editing
- THLP: investigation, writing – review & editing
- KNTT: investigation, writing – review & editing
- NTV: investigation, writing – review & editing
- NTXC: investigation, writing – review & editing
- VTT: investigation, writing – review & editing
- LPK: methodology, writing – review & editing
- CTT: investigation, writing – review & editing
- NTP: investigation, writing – review & editing
- NTHL: investigation, writing – review & editing
- LTHT: investigation, writing – review & editing
- NTCH: investigation, writing – review & editing
- PVT: investigation, writing – review & editing
- PTQ: investigation, writing – review & editing
- HTT: investigation, writing – review & editing
- NVH: investigation, writing – review & editing
- TVD: investigation, writing – review & editing
- EJ: investigation, writing – review & editing
- LR: investigation, writing – review & editing
- RBG: methodology, supervision, writing – review & editing
- HQC: investigation, methodology, writing – review & editing
- SY: conceptulalization, funding acquisition, methodology, supervision, writing – review & editing

## Supporting information

**S1 Table. Effect of overweight/obesity on full blood count and biochemistry.** Due to the non-linear trend of illness day in the models, the MDs (95% CIs) of overweight/obese group over healthy-weight group are derived from the linear mixed-effects models at some particular timepoints to present the differences between the two groups. P values are derived from the linear mixed-effects model. P overall represents the overall effect of overweight/obesity, and P trend compares the trend over time between the two groups. Abbreviation: ALT, alanine aminotransferase; AST, aspartate aminotransferase; CI, confidence interval; HDL-c, high density lipoprotein cholesterol; LDL-c, low density lipoprotein cholesterol; MD, mean difference

**S2 Table. Effect of overweight/obesity on endothelial, inflammatory, and lipid biomarkers.** The MDs (95% CIs) of overweight/obese group over healthy-weight group are derived from the linear mixed-effects models at some specific timepoints used in the models to present the differences between the two groups. P values are derived from the linear mixed-effects model. P overall represents the overall effect of overweight/obesity, and P trend compares the trend over time between the two groups. Abbreviations: CI, confidence interval; CRP, C-reactive protein; CXCL-10 (IP-10), CXC motif chemokine ligand 10; FU, follow-up; GDF-15, growth/differentiation factor 15; IL-10, interleukin-10; IL-18, interleukin-18; IL-1RA, interleukin-1 receptor antagonist; IL-6, interleukin-6; IL-8, interleukin-8; MD, mean difference; MPO, myeloperoxidase; NS1, nonstructural protein 1; PCSK9, proprotein convertase subtilisin/kexin type 9; VCAM-1, vascular cell adhesion molecule 1

**S3 Table. STROBE Statement—Checklist of items that should be included in reports of *cohort studies*.**

## Notes

### Funding Statement

Yes

### Author Declarations

The study was approved by the Ethics Committee of the Hospital for Tropical Diseases - Ho Chi Minh City - Vietnam (CS/BND/19/07) and the Oxford Tropical Research Ethics Committee (OxTREC 5-19)

